# Epigenetic Signatures of Asthma: A Comprehensive Study of DNA Methylation and Clinical Markers

**DOI:** 10.1101/2024.07.22.24310829

**Authors:** Austin J. Van Asselt, Jeffrey J. Beck, Brandon N. Johnson, Casey T. Finnicum, Noah Kallsen, Sarah Viet, Patricia Huizenga, Lannie Ligthart, Jouke-Jan Hottenga, René Pool, A.H Maitland-van der Zee, S.J. Vijverberg, Eco de Geus, Dorret I. Boomsma, Erik A. Ehli, Jenny van Dongen

## Abstract

**Background:** Asthma, a complex respiratory disease, presents with inflammatory symptoms in the lungs, blood, and other tissues. We investigated the relationship between DNA methylation and 35 clinical markers of asthma. The Illumina Infinium EPIC v1 methylation array was used to evaluate 742,442 CpGs in whole blood samples from 319 participants. They were part of the Netherlands Twin Register from families with at least one member suffering from severe asthma. Repeat blood samples were taken after 10 years from 182 of these individuals. Principal component analysis (PCA) on the clinical markers yielded ten principal components (PCs) that explained 92.8% of the total variance. We performed epigenome-wide association studies (EWAS) for each of the ten PCs correcting for familial structure and other covariates.

**Results:** 221 unique CpGs reached genome-wide significance at timepoint 1 (T1) after Bonferroni correction. PC7 accounted for the majority of associations (204), which correlated with loadings of eosinophil counts and immunoglobulin levels. Enrichment analysis via the EWAS Atlas identified 190 of these CpGs to be previously identified in EWASs of asthma and asthma-related traits. Proximity assessment to previously identified SNPs associated with asthma identified 17 unique SNPs within 1 MB of two of the 221 CpGs. EWAS in 182 individuals with epigenetic data at a second timepoint (T2) identified 49 significant CpGs. EWAS Atlas enrichment analysis indicated that 4 of the 49 were previously associated with asthma or asthma-related traits. Comparing the estimates of all the significant associations identified across the two time points (271 in total) yielded a correlation of 0.81.

**Conclusion:** We identified 270 unique CpGs that were associated with PC scores generated from 35 clinical markers of asthma, either cross-sectionally or 10 years later. A strong correlation was present between effect sizes at the 2 timepoints. Most associations were identified for PC7, which captured blood eosinophil counts and immunoglobulin levels and many of these CpGs have previous associations in earlier studies of asthma and asthma-related traits. The results point to using this robust DNA methylation profile as a new, stable biomarker for asthma.

## Introduction

Asthma affects approximately 262 million individuals worldwide and poses a significant health burden resulting in over 450,000 deaths annually [1, 2]. Often, the challenges associated with treating and diagnosing asthma stem from the complex nature of the disease [3] due to its multi-level heterogeneity with various clinical presentations, treatment responses, and disease trajectories [4]. Clinically, diagnosing asthma is done through the collection of several different clinical measurements and symptoms including a medical history assessment, a symptom and physical assessment, lung function tests, bronchial challenge tests, response to bronchodilators, and allergy testing [5]. These different diagnostic criteria, along with different individual phenotype characteristics, such as age of onset, have led to the identification of subtypes of asthma [6–8] and different endotypes with varying molecular underpinnings of the disease [6].

Several large-scale genomic studies have elucidated associations in multiple immune and regulatory genes for these endotypes, which may form the basis for polygenic scores [9]. Two of the largest genome-wide association studies (GWAS) of asthma were performed by Demenais et al. and Ferreira et al. The GWAS of asthma by Demenais et al. in 2018 included over 140,000 individuals and identified five novel, independent associations and confirmed several others that had previously been identified [10]. The study performed by Ferreira et al. in 2019, included over 300,000 individuals, identified 123 single nucleotide polymorphisms (SNPs) significantly associated with childhood asthma, 56 SNPs associated with adult-onset asthma, and 37 to be associated with both [11]. Collectively, Ferreira et. al. found 28 novel, independent associations between the two tested age groups [11]. Many of the associations that have been identified appear to be strongly linked to immune-related mechanisms [10, 11]. Though significant genetic associations were identified in both instances, the amount of total variance in relation to asthma they explained was limited (3.5-5.1%) [10, 11]. These outcomes reaffirm the notion that asthma, as a disease, is heavily influenced by environmental stimuli, which has led researchers to explore additional fields of study such as epigenetics and, more specifically, DNA methylation [12–14].

DNA methylation, as a molecular mechanism in humans, involves the addition of a methyl group to cytosine nucleotides, often in response to certain environmental queues [15–17]. The addition and removal of DNA methylation, specifically in regulatory regions of DNA, can lead to changes in gene expression and downstream cellular and tissue function [16, 17]. This entanglement with the external environment provides a basis for investigating the association of DNA methylation and asthma. The first studies of asthma and DNA methylation were conducted on candidate genes through techniques such as bisulfite pyrosequencing, which yielded moderate success highlighting CpG sites near *IFNy* as potential mediators of asthma [18, 19]. The candidate gene studies were followed by epigenome-wide association studies (EWAS) of asthma in different age cohorts, asthma subtypes, and tissue types [9], with most studies focusing on childhood, allergic asthma in whole blood and nasal epithelial samples [12, 13]. Studies of childhood asthma in nasal epithelial samples have identified several CpGs in both immune regulatory pathways and pathways of basic cell function [20–22]. Furthermore, a small number have shown to replicate in EWASs of whole blood, indicating some pan-tissue effects, though this effect appears to be limited [9]. Studies in adults found CpGs in genes relating to general inflammation, but the results show more variability in the CpGs that have been identified [23]. Though the majority of these results point towards immune-mediated pathways, the complete biological dynamic between these sites and their influence on asthmatic phenotypes has yet to be investigated [24]. Additionally, little is known regarding which measurable clinical markers associated with asthma may be contributing to these significantly associated CpGs, an area we explore here.

More generally, the continued identification of genetic and epigenetic variants influencing asthma is valuable for personalized treatment approaches. Here, we examined the genome-wide, CpG-specific methylation association with 35 different clinical markers of asthma from whole blood samples measured on the Illumina EPIC v1 methylation array and assessed the proximity of significantly associated CpGs to SNPs previously associated with asthma. We included individuals from families enrolled in the Netherlands Twin Register where at least one member of a nuclear family (consisting of 4 or 5 individuals) was diagnosed with asthma. Blood samples were collected at two timepoints, with 341 individuals having a sample collected at the first timepoint in the 1990’s and 233 individuals a second timepoint approximately 10 years later in the 2000’s [25]. Clinical markers were assessed at timepoint 1, while epigenetic data was generated on DNA from blood specimens for both timepoint 1 and timepoint 2.

## Results

### Clinical marker summary and data reduction

An initial set of 39 asthma-related clinical markers were measured. Markers that exhibited no variation between individuals were excluded (4 out of 39). Markers included measurements of lung function/capacity, skin prick test results measuring the reaction to common allergens, and others previously shown to be associated with asthma. Table 1 provides descriptions of the remaining 35 clinical markers. We performed an imputation step via the R package mice (v3.16.0) to supplement 12 of the 35 markers that had missing values. Distribution plots of these markers before and after imputation can be found in Supplemental Figure 1. We then performed principal component analysis on the 35 variables to reduce the data dimensionally using the r function robPCA (rospca version 1.0.4), which is specifically suited for non-normally distributed data [26]. The distributions of the principal component scores can be found in Supplemental Figure 2.

**Table 1.**
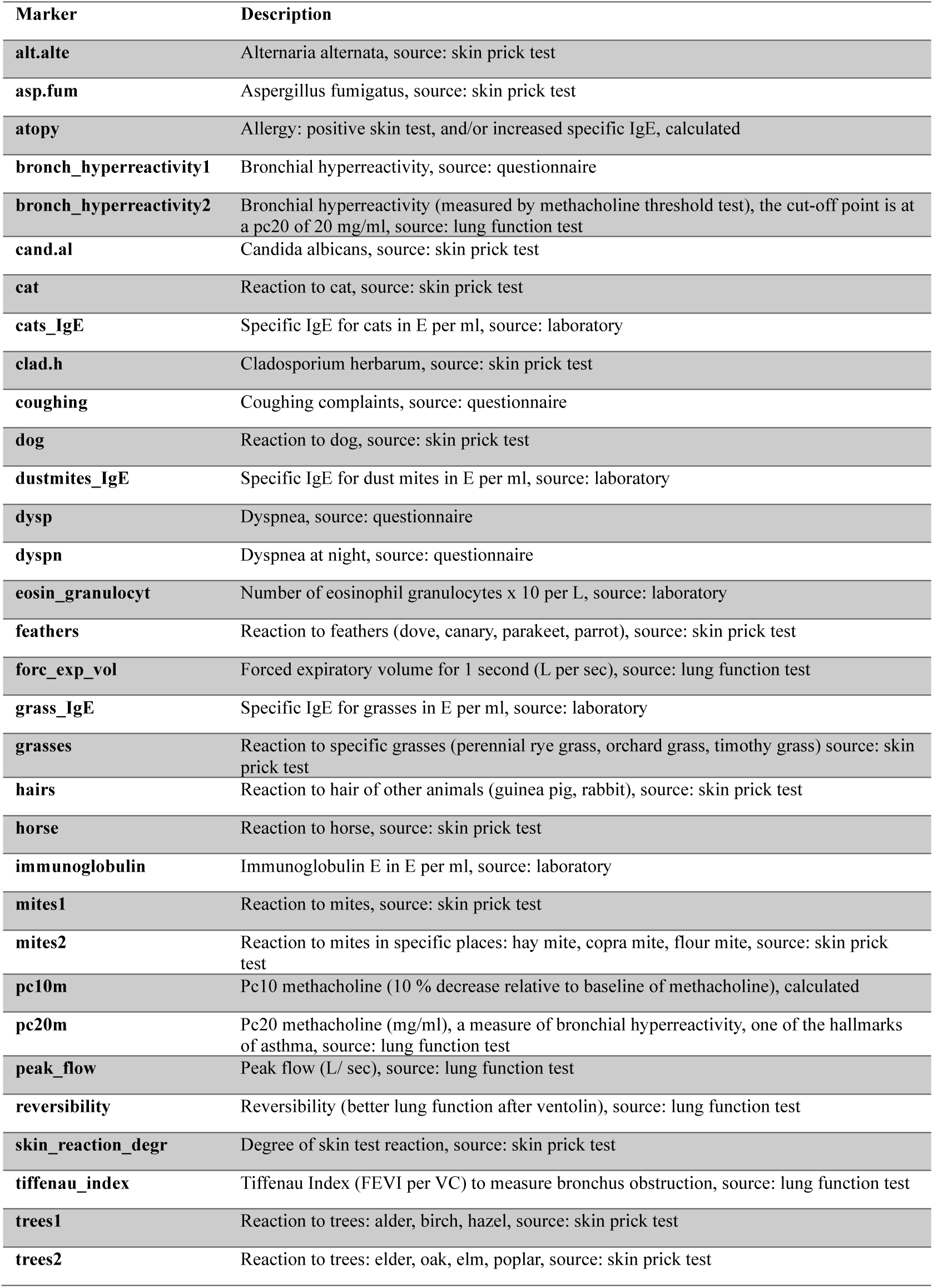
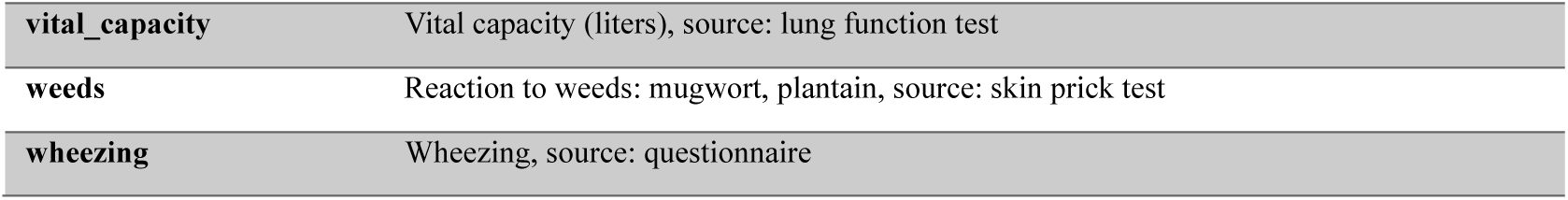
Descriptions/definitions for each clinical marker that was measured at T1.

| Marker | Description |
| --- | --- |
| alt.alte | Alternaria alternata, source: skin prick test |
| asp.fum | Aspergillus fumigatus, source: skin prick test |
| atopy | Allergy: positive skin test, and/or increased specific IgE, calculated |
| bronch_hyperreactivity1 | Bronchial hyperreactivity, source: questionnaire |
| bronch_hyperreactivity2 | Bronchial hyperreactivity (measured by methacholine threshold test), the cut-off point is at a pc20 of 20 mg/ml, source: lung function test |
| cand.al | Candida albicans, source: skin prick test |
| cat | Reaction to cat, source: skin prick test |
| cats_IgE | Specific IgE for cats in E per ml, source: laboratory |
| clad.h | Cladosporium herbarum, source: skin prick test |
| coughing | Coughing complaints, source: questionnaire |
| dog | Reaction to dog, source: skin prick test |
| dustmites_IgE | Specific IgE for dust mites in E per ml, source: laboratory |
| dysp | Dyspnea, source: questionnaire |
| dyspn | Dyspnea at night, source: questionnaire |
| eosin_granulocyt | Number of eosinophil granulocytes x 10 per L, source: laboratory |
| feathers | Reaction to feathers (dove, canary, parakeet, parrot), source: skin prick test |
| forc_exp_vol | Forced expiratory volume for 1 second (L per sec), source: lung function test |
| grass_IgE | Specific IgE for grasses in E per ml, source: laboratory |
| grasses | Reaction to specific grasses (perennial rye grass, orchard grass, timothy grass) source: skin prick test |
| hairs | Reaction to hair of other animals (guinea pig, rabbit), source: skin prick test |
| horse | Reaction to horse, source: skin prick test |
| immunoglobulin | Immunoglobulin E in E per ml, source: laboratory |
| mites1 | Reaction to mites, source: skin prick test |
| mites2 | Reaction to mites in specific places: hay mite, copra mite, flour mite, source: skin prick test |
| pc10m | Pc10 methacholine (10 % decrease relative to baseline of methacholine), calculated |
| pc20m | Pc20 methacholine (mg/ml), a measure of bronchial hyperreactivity, one of the hallmarks of asthma, source: lung function test |
| peak_flow | Peak flow (L/ sec), source: lung function test |
| reversibility | Reversibility (better lung function after ventolin), source: lung function test |
| skin_reaction_degr | Degree of skin test reaction, source: skin prick test |
| tiffenau_index | Tiffenau Index (FEV1 per VC) to measure bronchus obstruction, source: lung function test |
| trees1 | Reaction to trees: alder, birch, hazel, source: skin prick test |
| trees2 | Reaction to trees: elder, oak, elm, poplar, source: skin prick test |
| **vital_capacity** | Vital capacity (liters), source: lung function test |
| **weeds** | Reaction to weeds: mugwort, plantain, source: skin prick test |
| **wheezing** | Wheezing, source: questionnaire |

PCA investigating the multidimensional structure of the clinical marker data showed the first principal component to account for 35.27% of the variance. Principal component (PC) 1 was largely associated with methacholine challenge values, other measurements of lung capacity/functionality, and lung ailments such as coughing and wheezing. PCs 2, 3, and 4 accounted for 19.54%, 12.46%, and 9.75% of the variance, respectively, and were also mostly associated with functional lung measurements. PCs 5 and 6 accounted for 8.39% and 6.05% of the variance, respectively, and showed strong association with the FEV1/FVC ratio, also called modified Tiffeneau-Pinelli index. PC7 accounted for 4.28% of the variance and showed very strong associations to eosinophil counts and immunoglobulin levels. PCs 8, 9 and 10 accounted for 2.20%, 1.22%, and 0.84% of the variance and each associated with a variety of allergen response measurements (Figure 1). Supplemental Table 1 contains the raw eigen values and percentage of variance explained for each PC.

**Figure 1.**
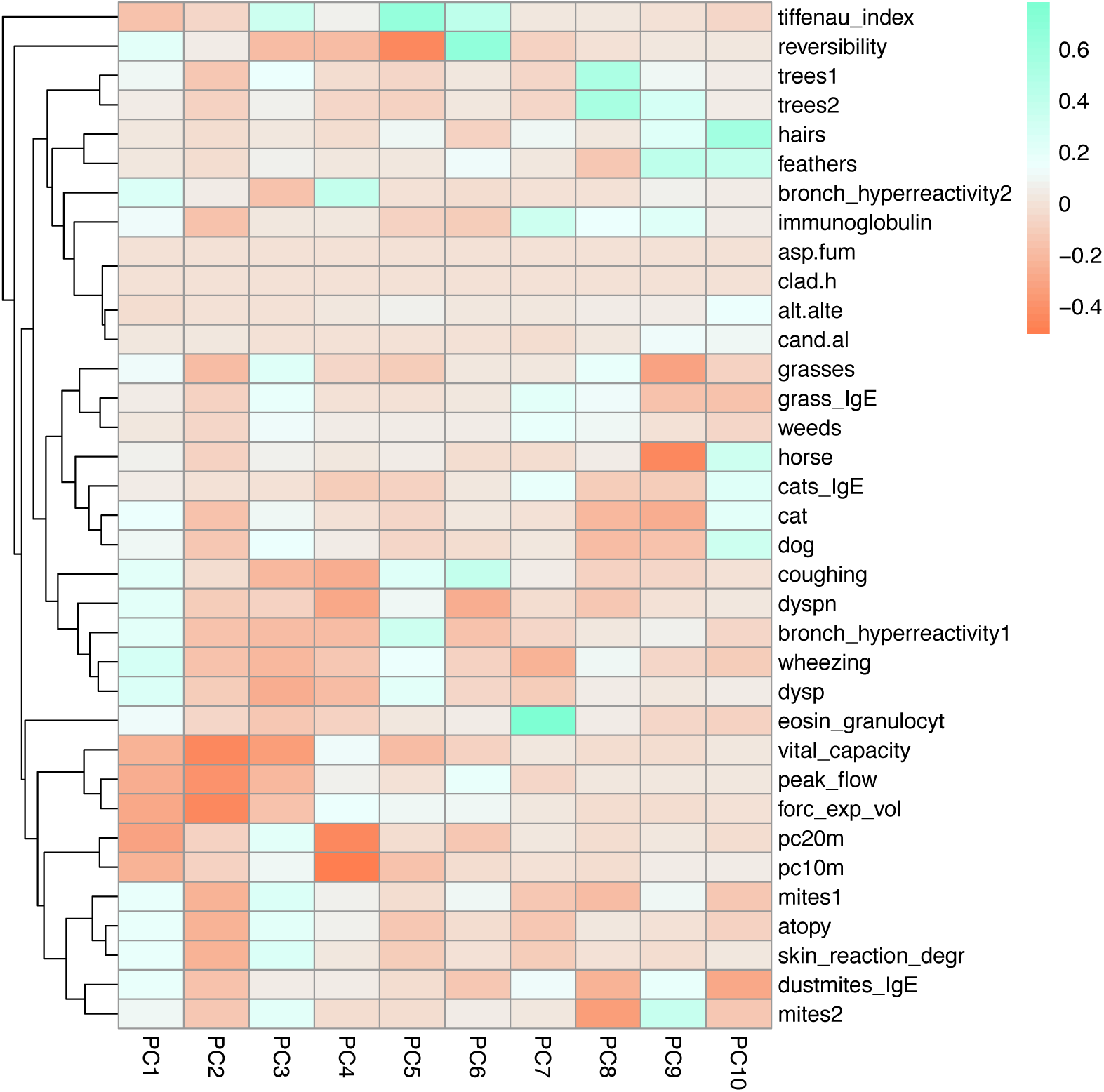
Heatmap showing correlations between the clinical asthma markers and principal components generated from the asthma marker data. An explanation of the variables is provided in Table 1.

### Timepoint 1 EWAS via 10 Generated PCs

Epigenome-wide association studies of each of the ten principal components yielded multiple CpGs that reached genome-wide significance. In total, we identified 222 significantly associated CpGs (following a Bonferroni correction, α = 0.05 / 742,442). Of the 222, one, cg07329820, was associated with two PCs (PCs 9 and 10), resulting in a total of 221 unique CpGs that were significantly associated. Table 2 provides a summary of the covariate frequencies for the sample population. Table 3 summarizes the number of CpGs identified in each EWAS and Supplemental Figure 3 shows the results of each individual EWAS via Manhattan plots. Inflation values for each EWAS can be found in Supplemental Table 2. A complete list of all the CpGs significantly associated at T1 can be found in Supplemental Table 3, and complete summary statistics for each of the EWASs can be found in Supplemental Table 4.

**Table 2.**
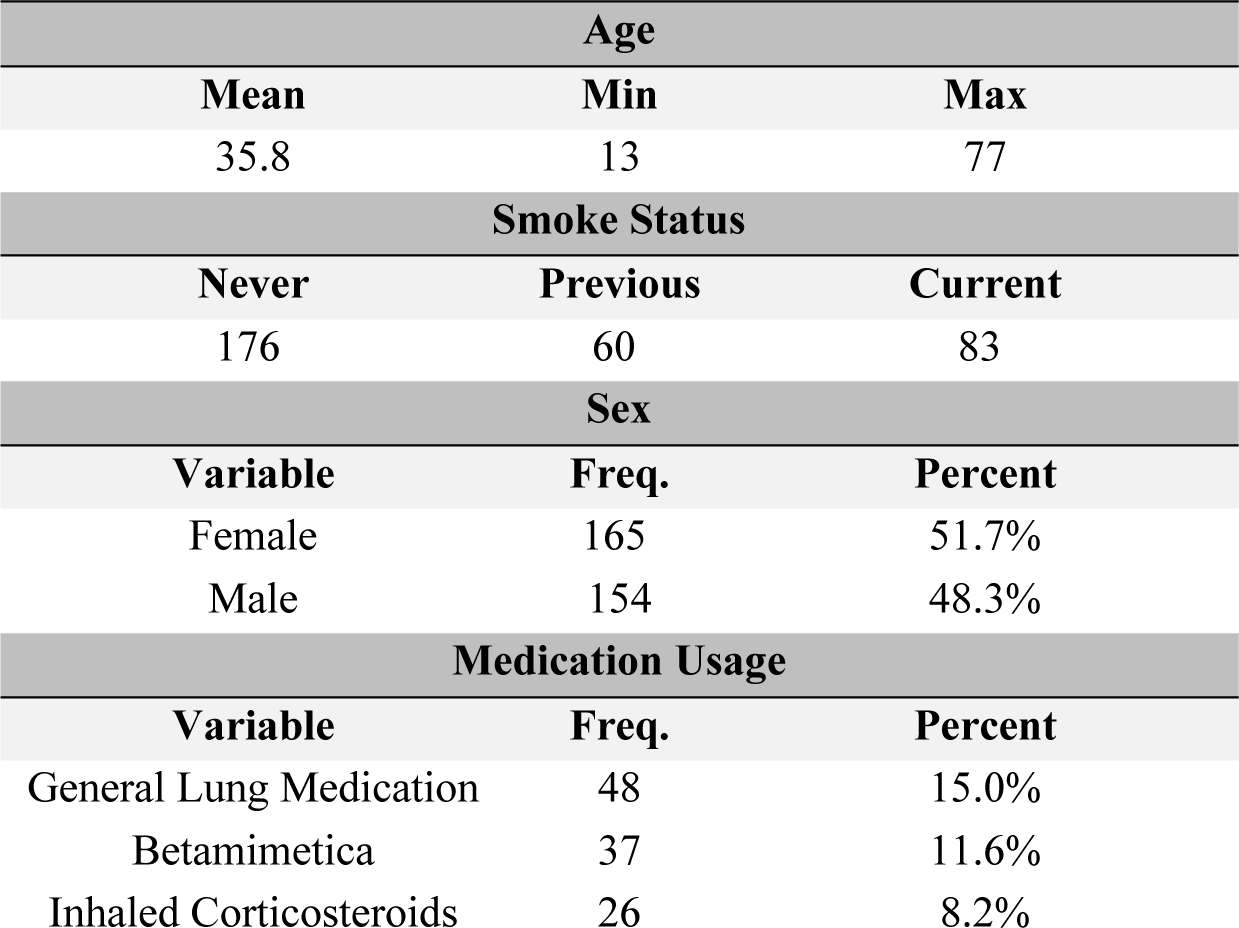
Sample population covariate frequencies.

| Age |  |  |
| --- | --- | --- |
| Mean | Min | Max |
| 35.8 | 13 | 77 |
| Smoke Status |  |  |
| Never | Previous | Current |
| 176 | 60 | 83 |
| Sex |  |  |
| Variable | Freq. | Percent |
| Female | 165 | 51.7% |
| Male | 154 | 48.3% |
| Medication Usage |  |  |
| Variable | Freq. | Percent |
| General Lung Medication | 48 | 15.0% |
| Betamimetica | 37 | 11.6% |
| Inhaled Corticosteroids | 26 | 8.2% |

**Table 3.**
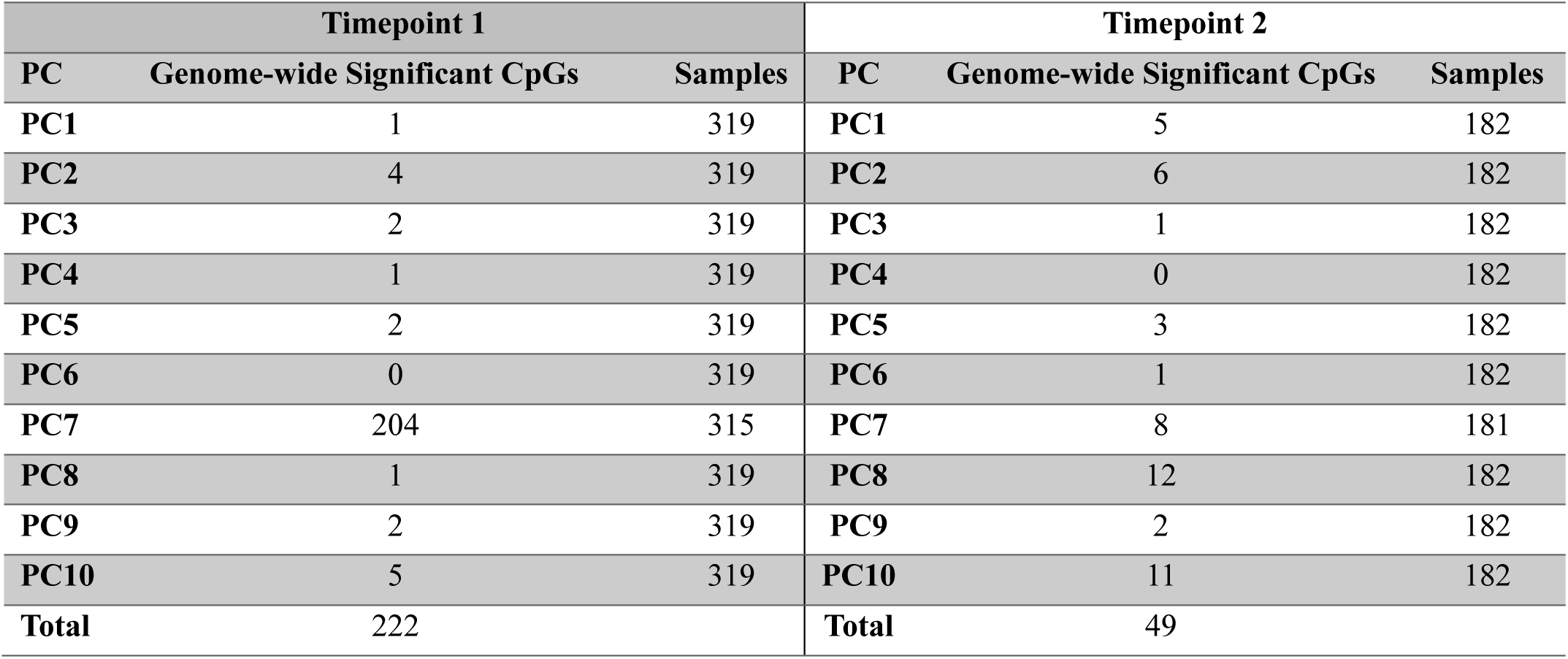
A summary of the genome-wide significant CpGs identified in each EWAS for time point 1 and timepoint 2.

| Timepoint 1 |  |  | Timepoint 2 |  |  |
| --- | --- | --- | --- | --- | --- |
| PC | Genome-wide Significant CpGs | Samples | PC | Genome-wide Significant CpGs | Samples |
| PC1 | 1 | 319 | PC1 | 5 | 182 |
| PC2 | 4 | 319 | PC2 | 6 | 182 |
| PC3 | 2 | 319 | PC3 | 1 | 182 |
| PC4 | 1 | 319 | PC4 | 0 | 182 |
| PC5 | 2 | 319 | PC5 | 3 | 182 |
| PC6 | 0 | 319 | PC6 | 1 | 182 |
| PC7 | 204 | 315 | PC7 | 8 | 181 |
| PC8 | 1 | 319 | PC8 | 12 | 182 |
| PC9 | 2 | 319 | PC9 | 2 | 182 |
| PC10 | 5 | 319 | PC10 | 11 | 182 |
| Total | 222 |  | Total | 49 |  |

By far, the largest number of CpGs were associated with PC7 (which mostly captures variance from eosinophil counts and immunoglobulin levels). These 204 CpGs, largely, are composed of CpGs with a high average methylation level and almost exclusively show negative effect sizes, such that higher scores on PC7 correlate with lower methylation levels. The PC10 EWAS identified associations with 5 CpGs, followed by the PC2 EWAS with 4 significantly associated CpGs. EWASs for PCs 3, 5, and 9 showed significant associations with 2 CpGs. EWASs for PCs 1, 4, and 8 were significantly associated with 1 CpG, while the EWAS for PC6 was the only association study that did not identify any significantly associated CpGs.

An enrichment analysis via the online EWAS Atlas (https://ngdc.cncb.ac.cn/ewas/atlas/index, accessed June 11, 2024) for the 221 unique CpGs was performed. This analysis identified that 71 (32.1%) and 173 (78.3%) of these CpGs had previously been identified to associate with allergic asthma and fractional exhaled nitric oxide (FENO), respectively (shown in Figure 2). Further, asthma related phenotypes such as childhood asthma, allergic sensitization, atopy, serum IgE levels, and others were shown to be previously associated with CpGs identified in our analysis. In total, 190 of the 221 CpGs (86.0%) were shown to have a previously identified association with asthma or some asthma marker, leaving 31 novel, unique associations. This enrichment analysis also highlighted significant overlap with the FoxO signalling pathway (7 CpGs previously associated). Enrichment analysis restricting to the 204 CpGs associated with PC7 recapitulated these findings (Supplemental Figure 4), showing that all of the previously associated CpGs were identified via this PC7 EWAS. We then performed an additional query of the eFORGE online database (https://eforge.altiusinstitute.org/, accessed July 2^nd^, 2024) with the 204 CpGs associated with PC7 to identify overlap with cell-type specific DNA regulatory elements. This identified significant associations with regulatory elements in multiple tissue sources, with the greatest number of significant findings (31 in total) associating with various subpopulations of white blood cells (Supplemental Figure 5).

**Figure 2.**
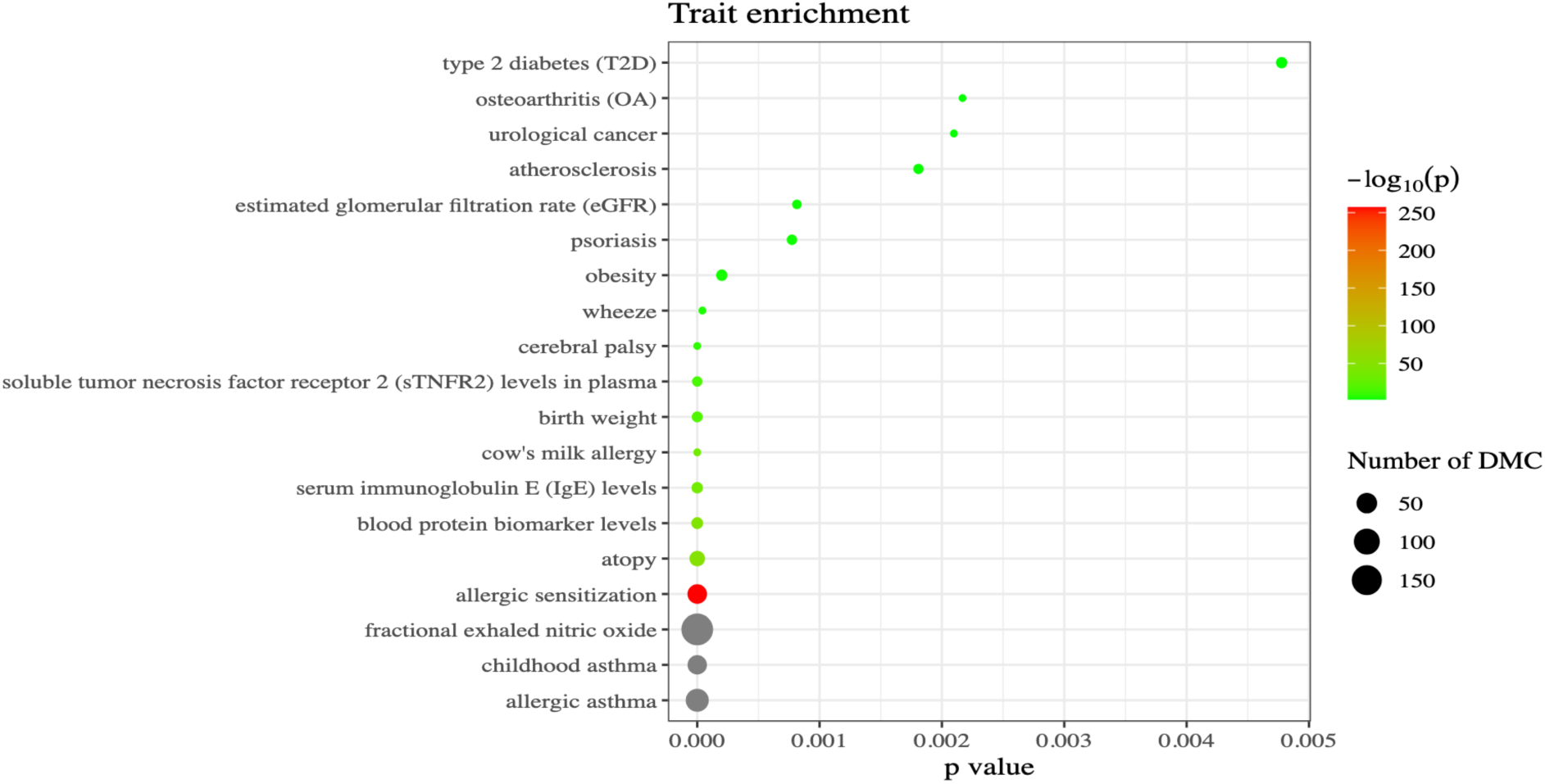
EWAS atlas enrichment analysis of the 221 unique CpGs identified at TP1 across the 10 EWASs. The plot shows the number of previously identified Differentially Methylated CpGs (DMC) that are present within the CpGs identified in our analysis. The EWAS Atlas job can be found via this job ID: ea5abe0e33f28b27ab3948e2ccd4044c.

### Analysis of overlap with previously identified SNPs

An investigation of proximity with the 221 unique significantly associated CpGs was performed to identify SNPs that have been previously identified by Demenais et al. to associate with asthma (p< 5 × 10^-8^, N= 892 SNPs). This assessment was performed by searching for SNPs within 1 MB upstream and downstream of each CpG. We identified a total of 17 unique SNPs (1.9% of all asthma-associated SNPs) to reside within 1 MB of a CpG identified at time point 1. These 17 unique SNPs involved only two unique CpGs (cg02046836, cg23515090). A summary of these SNPs within proximity to each CpG is in Supplemental Table 5. Additionally, a query of the BBMRI methylation Quantitative Trait Loci (mQTL) database with these two CpGs was performed, which showed that the asthma GWAS SNPs are not mQTLs for cg02046836, cg23515090, and that these CpGs are also not associated with any other SNPs [27].

### Timepoint 2 EWAS

Of the initial 319 individuals who were included at the first timepoint, 182 individuals had a second whole blood sample collected approximately 10 years later. The DNA from these blood samples were also profiled on Illumina EPIC v1 methylation arrays (but no detailed asthma phenotyping was performed at follow-up). We repeated the EWASs for the ten PCs with DNA methylation data at T2. The EWASs of the ten PCs with DNA methylation data from samples collected ten years after asthma phenotyping identified 49 unique CpGs at epigenome-wide significance. While none of the CpGs identified at T1 were significant at T2, three CpGs identified at T2 were within 100 kb of a significant CpG identified at timepoint 1 (Supplemental Table 8). Furthermore, we assessed the correlation (Pearson’s) of the estimates for all 271 significant associations (222 at T1, 49 at T2) across the two time points, and we identified a strong correlation (r = 0.82). Table 3 outlines the number of CpGs identified in each time point 2 EWAS and Supplemental Figure 6 contains the Manhattan plots for these EWASs. Supplemental Table 6 contains all of the 49 significantly associated CpGs identified at T2, and Supplemental Table 7 states the inflation values for each of these 10 EWASs.

## Discussion

Asthma is a complex chronic disease that has been shown to be influenced by multiple factors including genetics and epigenetics. Here, we have identified 221 unique CpGs that significantly associate with PC scores generated from an advanced, comprehensive panel of 35 clinical markers of asthma. Of these, 190 have previously been identified to associate with asthma and asthma-related phenotypes, while the other 31 CpGs are novel findings. Further, two of these CpGs reside within close proximity (1 MB) to 17 SNPs previously identified in a large GWAS of asthma (source). We then performed a longitudinal assessment using methylation data from whole blood samples collected approximately 10 years and identified 49 unique significantly associated CpGs. Finally, we yielded a correlation of the estimates across the two timepoints of 0.82 from the 271 (222 T1 + 49 T2) significant associations.

Historically, asthma as a disease has been revealed to be extremely challenging to identify the root cause of and treat effectively [28]. The disease heterogeneity and mixed involvement of environmental stimuli and the genome have largely contributed to this difficulty. To avoid the complications that can come with this, we utilized measured clinical markers of asthma in young adults and their middle-aged parents from families with a history of asthma burden where at least one individual within the family had been diagnosed with asthma in some capacity. For each individual, an extensive number of clinical markers related to asthma were captured. This comprehensive dataset, while immensely valuable with its depth, is too overwhelming to glean the valuable nuances through individual assessments of each specific marker. Additionally, some markers that were collected lacked a normal distribution, which could reduce their performance in a traditional EWAS. For this reason, we utilized PCA to simplify the 35 different variables, and their accompanying variance, into 10 PCs with each successive PC capturing a lesser and different portion of the overall variance within the data set. This method reduced the total number of variables to investigate from 35 to 10, making comparisons of it with a rich DNA methylation dataset much more feasible. Additionally, the PCs maintained a much more normal distribution, making them ideal for EWAS. This method of phenotype data reduction and handling has been previously implemented in a GWAS of rice variants, where GWASs were successfully performed using the PC scores from multiple phenotype variables [29]. To our knowledge, this is the first instance of implementing it for epigenome-wide association studies.

Through our EWASs of each of the 10 PCs, we identified 222 CpGs to be significantly associated with one of the PCs derived from the 35 clinical asthma markers. Of these 222 associations, only a single CpG was found to be replicated across multiple principal components, leaving 221 unique CpGs to be significantly associated. The single overlapping CPG, cg07329820, was identified with PCs 9 and 10. These two PCs align with asthma markers that are very similar (largely allergen response measures), which could explain why they identified the same CpG.

A breakdown of the identified CpGs from each PC shows some intriguing details. PC7 had the greatest number of CpG associations with 204. While this PC accounts for only 4.3% of the total clinical marker variance, it almost exclusively represents the variance associated with blood eosinophil counts and blood immunoglobulin levels. This large number of associations could be indicative of the eosinophilic inflammatory response typically seen in allergic asthma. Previous literature has shown that methylation experiments in whole blood may capture more of the Th2-mediated immune response due to the nature of the cell population, making it the ideal sample source for identifying this type of association [30–32]. It is critical to note that because the proportions of cells themselves in a sample can drastically influence the measurement of methylation, we made sure to incorporate cell proportion estimates as a covariate in our analyses. Without this inclusion, it could be argued that the findings identified here are only due to differences in cellular composition. With this correction in mind, it may be the case that the differential methylation identified here is something global, across multiple immune cell types, either in response to or to promote the eosinophilic inflammatory pathway. A query of the EWAS Atlas of these 204 CpGs showed that, almost exclusively, the CpGs we identified have a previous association with asthma and measurements of asthma (Supplemental Figure 4). In total 190 of the 204 CpGs were shown to have some form of previous association (93.1%), leaving 14 novel associations. Since the majority of overlap of our findings with previous EWASs of asthma, asthma phenotypes, and symptoms are captured within this EWAS of PC7, we postulate that many of the previous findings generated in studies utilizing whole blood samples could be identifying associations with the underlying influences of increased eosinophil counts/activation and increased immunoglobulin levels.

An additional investigation into these 204 CpGs via the eFORGE online database identified multiple significantly associated tissue-specific regulatory elements (Supplemental Figure 5). The greatest number of associations were from subpopulations of white blood cells in enhancers and weak transcriptional activators. This is to be expected considering our results were generated from samples of whole blood. Many of the subpopulations identified here are lymphocytes, potentially indicating that the methylation profiles of these adaptive immune cells may be altered due to the changes in eosinophil counts and immunoglobulin levels. The tissue group with the next largest number of significant associations was the digestive tract, with six significant associations to various subpopulations of cells. These associations could hint at the conserved role that all mucosal and epithelial barriers play in pathogen protection and the body’s response to the external environment.

A KEGG pathway enrichment analysis (conducted via the EWAS Atlas) of the 221 unique CpGs highlighted the FoxO signaling pathway, showing that 7 of the CpGs (all found within the PC7 EWAS) have been previously identified to associate with this pathway. This pathway is of particular interest due its significant role in regulating apoptosis, glucose metabolism, oxidative stress, and longevity, which has led to findings associating its dysregulation with other illnesses such as Alzheimer’s disease, type 2 diabetes, and cancer [33, 34]. It has been previously shown that FoxO1 modulates IL-9 generating Th9 cells [35]. Further, overexpression of IL-9 in the lungs can result eosinophilic inflammatory infiltration and mucus secretion [35, 36]. Additionally, FoxO3 has been found to be expressed in airway epithelium playing a critical role in controlling the innate immune response to airway infections [36]. It could be the case, based on these intertangled findings of DNA methylation with eosinophilic presence and FoxO signaling, that epigenetic modulation of FoxO signaling plays a role in its contribution to asthma and airway inflammation.

Principal component 1 identified a single significantly associated CpG, cg18182148. While this principal component does capture the largest amount of variance (35.3%), the variance it captures is largely from one specific subset of measurements, most notably those measuring general lung functionality such as vital capacity, peak flow, forced expiratory volume, and methacholine challenge response. This is continued with PC2, PC3, and PC4 which were significantly associated with 4, 2, and 1 CpGs, respectively. These PCs, like PC1, capture a significant amount of variance from these same lung function measurements. This may indicate that these measurements of lung function, though important in characterizing symptoms of asthma, may have limited identifiable associations with DNA methylation. The specific CpG significantly associated with PC1, cg18182148, is the only CpG of these identified here related to immune regulation. It has not been previously shown to associate with asthma phenotypes, though it has been previously identified in studies of various cancers of the prostate, liver, and colon (via the EWAS Atlas) [37–40]. These associations are likely due to its placement within the transcription start site (TSS) of GFI-1, a strong oncogenic and hematopoietic regulator. GFI-1 plays a critical role in lymphoid differentiation, which could be the reason for its association here due to the large influence the immune system can play on lung functionality and, more generally, asthma as a whole [41]. While the use of a whole blood sample, as done here, is logical when investigating an immune-mediated illness such as asthma, an investigation of these lung function measurements in association with DNA methylation measured via a lung epithelial sample may highlight additional findings not seen here and provide more information on the local cellular environment responsible for the specific differences in lung function.

Principal components 5 and 6 capture variance associated with bronchial hyperreactivity, coughing, reversibility (of lung function following the administering of ventolin), and Tiffenau Index. PC5 was significantly associated with 2 CpGs, while PC6 did not have any significant associations. The two CpGs identified here, cg07454584 and cg18561513, have not been previously shown to have any associations with asthma or other immune-mediated illnesses (via the EWAS Atlas). Like the measurements captured in PCs 1-4, these asthma markers, while important for asthma diagnosis and the understanding of symptoms, may not have large quantities of associations with DNA methylation measured via whole blood.

Principal components 8, 9, and 10 contain much of the variance from the numerous allergen tests included in this study (containing both skin prick test results and serum specific IgE measurements). These PCs were significantly associated with one, two, and 5 CpGs, respectively. Of the 8 CpGs identified here, all but one has not been previously shown to associate with chronic illnesses or immune-mediated diseases. Cg14161241, which was significantly associated with PC10, is the lone exception with previous associations with obesity and type 2 diabetes [42]. Like many of the other markers represented via PCs 1-6, the effect on DNA methylation measurable via a whole blood sample is likely limited with these markers.

Due to the well-documented genetic contributions to asthma, we next wanted to compare our findings within the methylome to those previously discovered in the genome via large GWASs [10, 43]. Utilizing the findings of Demenais et al., we searched for significantly associated SNPs within 1 MB of the 221 unique CpGs that we identified. Of the significantly associated SNPs (p< 5 × 10^-8^, N= 892 SNPs), we found 17 (1.9%) to be within 1 MB of 2 of the 221 CpGs that we identified (cg02046836, cg23515090). Interestingly, neither of these two CpGs have been previously identified as an mQTL [27]. The proximity of these SNPs to the identified CpGs could be caused from multiple factors outside of known standard mQTL associations. It could be the case that these CpGs do have some genetic influence acting on their methylation status that has not yet been characterized. Additionally, it is certainly possible that the SNPs and CpGs being within close proximity to one another is simply a random occurrence, and their presence and downstream effects are independent of one another. To summarize, we found limited overlap of our EWAS signal with top loci from a previously performed GWAS of asthma and that these significant CpGs nearby these GWAS SNPs were not directly affected by mQTLs, which suggests that our EWAS largely captures independent epigenetic signal at loci that also harbor small amounts of genetic variants that influence genetic susceptibility to asthma.

Thus far, we have shown significant associations with clinical markers of asthma from a singular, cross-sectional timepoint that corroborates other previous findings from similar studies. However, a potential increase in our understanding of asthma may lie in investigating the disease to some longitudinal capacity. Two previous studies in the Isle of Wight Birth Cohort (IOWBC) were conducted in 2022, which investigated the association of DNA methylation with asthma acquisition across adolescence and adulthood [44, 45]. In each of these studies, several CpGs, with some residing in immune regulatory genes, were identified to be associated with asthma acquisition [44, 45]. The findings from these two studies highlight promising insight into the potential longitudinal persistence of DNA methylation signatures associated with asthma, though additional studies are needed to investigate this persistence later in life (adulthood), in additional cohorts, and how they relate to specific measurements of asthma.

After our initial findings, with the availability of an additional sample collected significantly later for many of the individuals, we were interested in investigating the longitudinal persistence of the DNA methylation signature that was identified to associate with asthma markers at the first timepoint. This analysis was based on the same PC data from the original 35 clinical markers measured at baseline and DNA methylation at follow-up from a sample collected ∼10 years later. The results from these additional 10 EWASs showed very little overlap with the findings from the initial assessment. In total, these 10 EWASs identified a total of 49 significantly associated CpGs (of which none were replicated across multiple PCs). No CpGs were replicated across the two timepoints, however, when looking within a 100 kb window of proximity, we found that 3 CpGs identified at timepoint 2 (found via PCs 2, 3, and 7) were within 100 kb of 4 CpGs identified at T1 (all found via PC7). The low direct overlap between the two timepoints could be due to the methylation signature associating with asthma diminishing over time, potentially as individuals’ asthma status and severity changes. A query of the EWAS Atlas, however, using these 49 CpGs identified at T2 showed that 3 and 1 CpGs have been previously associated with FENO and allergic sensitization, respectively (Figure 3). This highlights that there may be some longitudinal persistence of an asthmatic epigenetic signature. Further, we identified, when comparing the estimates of all 271 significant associations (222 + 49) across the two timepoints, a strong correlation of 0.82 adding to the notion that some long-term persistence of the asthma associated methylation signature may be occurring. This consistency of the EWAS estimates indicates that replication of specific CpGs could occur with an increased sample size. Additional studies investigating the long-term lung functionality following asthma diagnosis, even in cases of remission, could benefit from investigating DNA methylation longitudinally in a more nuanced, complete manner.

**Figure 3.**
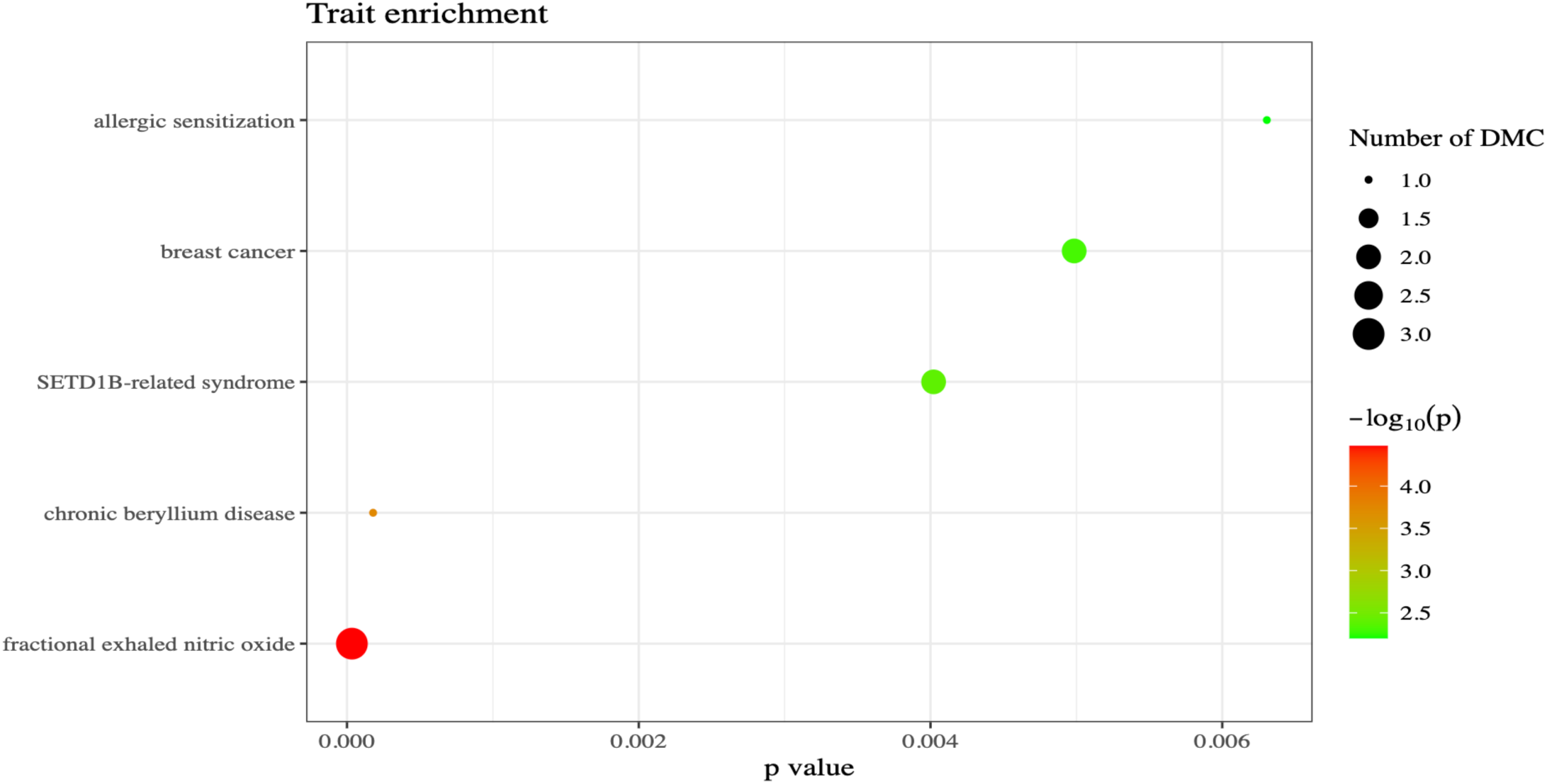
EWAS atlas enrichment analysis of the 49 unique CpGs identified at TP2 across the 10 EWASs. The plot shows the number of previously identified Differentially Methylated CpGs (DMC) that are present within the CpGs identified in our analysis. The EWAS Atlas job can be found via this job ID: a3e03507f39d7af4c7a82d0b0725308e.

Our study does have some limitations that should be considered. First, all individuals included in this study are of European descent and from the Netherlands. Asthma prevalence can differ greatly depending on the local environment of the individual and their ethnicity. Including a diverse population could improve the generality of the findings to other groups of individuals. Additionally, the results generated from the second time point contained a significantly reduced number of individuals due to sample availability, which could contribute to the differences reported. We also did not have clinical marker measurements available from the second timepoint, which could have shed some insight on the persistence of the asthma symptoms and severity at that time. Overall, increasing the number of individuals at both timepoints using multiple individuals from varying backgrounds would be the most optimal strategy and offer the most comprehensive assessment of asthma.

## Conclusion

Asthma, along with many other chronic, immune-mediated diseases, is challenging to diagnose, classify, and treat. Here, we contribute a comprehensive assessment of the relationship between DNA methylation and a collection of clinical asthma markers. We identified 270 unique, significant CpGs to be associated with principal component scores based on clinical asthma markers. The large majority of these CpGs were associated with PC7, which represented eosinophil counts and blood immunoglobulin levels and 190 were previously associated with asthma, asthma markers, and other allergic illnesses. We also investigated the proximity of these 221 identified unique CpGs to previously identified SNPs that associate with asthma. We found 17 unique SNPs that were previously identified to be associated with asthma within 1 MB of two of the 221 CpGs reported here demonstrating limited proximal overlap with these known genetic associations. Over approximately 10 years we showed that the correlation of the estimates across the two timepoints from the 271 significant associations was strong at 0.82, pointing to a persistent methylation signature associated with asthma. Thus, these results highlight a robust, persistent epigenetic signal in whole blood associated with asthma and, more specifically, a selection of clinical asthma markers.

## Methods

### Participants

The study participants and the enrollment process have been previously described [46–48]. Participants came from twin families (young adult twins and their parents) from the Netherlands [48]. To be eligible for the study, at least one member of the family had to report to have been diagnosed with asthma, indicated by self-report in the Netherlands Twin Register (NTR) survey 1 or 2, collected in the early 1990s (please see Supplementary Tables 9 and 10). During a visit to the Vrije Universiteit Medical Center (VUMC), blood samples and over 40 biological and clinical measures of asthma were collected from 425 persons [49]. A complete list of the clinical markers that were collected can be found in Table 1. A subset of 233 persons participated in the NTR Biobank project between 2004 and 2008 in which blood and buccal samples were collected [25]. DNA was extracted at the time of sample collection or immediately prior to the assessment of methylation [50]. All samples were simultaneously measured on the Illumina Infinium EPIC v1 methylation array. In total, 375 of the 425 individuals had a sample remaining for analysis, and 341 of these had good quality Illumina blood EPIC Array data from at least the first time point, and 232 individuals had blood Illumina EPIC Array data from two time points.

The study was approved by the Central Ethics Committee on Research Involving Human Subjects of the VU University Medical Centre, Amsterdam, an Institutional Review Board certified by the U.S. Office of Human Research Protections (IRB number IRB00002991 under Federal-wide Assurance-FWA00017598; IRB/institute codes, NTR 03-180).

### Sample collection and DNA extraction

The procedures of whole blood sample collection have been previously described [25, 46, 51]. The genomic material analyzed in this study have been extracted and assessed for DNA quantity, quality, purity, and individual identity. The DNA analyzed in this study was extracted from whole blood samples using the Zymo Quick DNA mini-prep Kit [50]. Genomic material was quantified via the Invitrogen Qubit Broad-Range Fluorescent Assay, and sample purity was assessed using standard absorbance metrics via a SpectraMax microplate reader.

### Genotyping

The genotyping process has been previously described [47, 52]. Briefly, genotype data was generated via the Illumina Infinium Global Screening Array. All sample data quality was assessed vigorously, and the refined genotype data was used to confirm familial relationships and sample sex. In total, 20 individuals were omitted for analysis due to mismatched familial identities.

### DNA Methylation Assessment

DNA bisulfite conversion was performed utilizing the Zymo EZ-96 DNA Methylation Kit [53]. DNA methylation was assessed using the Illumina Infinium EPIC v1 DNA Methylation Array on all samples at the Avera Genetics Laboratory [54]. The samples were fully randomized across arrays.

### DNA Methylation Data Quality Control

Quality control has been described in detail previously [47]. In brief, DNA methylation data quality was performed using two bioinformatic tools. First, DNA methylation data was preliminarily assessed using the Illumina GenomeStudio 1.0 software. Second, methylation data quality control and normalization were conducted using a pipeline developed by the Biobank-based Integrative Omics Study (BIOS) Consortium in R (version 4.4.0). Sample quality was first assessed using the R package MethylAid (v1.38.0) to omit any outliers in sample performance (default thresholds) [55]. Array probe filtering and functional normalization were performed using the R package DNAmArray (v2.2.0) [56]. The R package omicsPrint (v.1.24.0) was utilized to identify sample genotypes based on methylation probes to verify sample relationships gathered from genotype data via the GSA [57]. The functions getSex from DNAmArray and the R package meffil (v1.3.8) were used to identify and confirm sample sex based on X chromosome methylation pattern [58]. The following probe filters were applied: Probes were set to missing (NA) in a sample if they had an intensity value of exactly zero, detection P-value > 0.01, or bead count < 3. DNAmArray will also remove any probes that show a success rate below 0.95 across all samples. In total, this filtering process reduced our number of individuals to 341 (from the original 375). Finally, previous studies performed by Zhou et al. in 2017 identified polymorphic and cross-reactive probes that are included on Illumina methylation array platforms [59]. Generally, they recommend an omitting of approximately 100,000 probes on the EPIC methylation array. This removal process was carried out using the probemasking() function via DNAmArray. Following these steps, a total of 759,263 methylation sites were included in this study out of the total 865,859 possible CpGs. Only autosomal methylation sites were considered for downstream analyses, which left a total of 742,442 CpGs included in the final analyses. Finally, we calculated cellular proportions for all samples using the IDOL whole blood reference library [60].

### Covariates

Several covariates were utilized in our epigenome-wide association studies. For each sample at each timepoint, covariate data was collected at the same time. We included age, sex, array row, sample plate (dummy-coded), smoking status, asthma/lung medication use, and cellular proportions. For smoking status, individuals were classified into three groups (codes as 0 = Never smoker, 1 = former smoker, 2 = current smoker). Cellular proportions (natural killer cells, monocytes, B cells, CD4+ T cells, CD8+T cells) for each of the whole blood samples were estimated via the methylation data from that sample. The usage of specific and general asthma medications (inhaled corticosteroids (0=no, 1=yes), betamimetica (0=no, 1=yes), general lung medication (0=no, 1=yes)) was recorded for each participant at the first time point, and all three medications were included as covariates for the T1 EWAS. The information gathered here was used at both time points. Due to limited variability in medication use at the second timepoint, only a single “general lung medication use” variable was used as a covariate for T2.

### Methylation Data Annotation

Genomic annotations were gathered from the EPIC manifest file that is provided by Illumina (MethylationEPIC_v-1-0_B5.csv): locations of CpG islands, ENCODE DNase I Hypersensitive sites (DHSs), ENCODE transcription factor binding sites (TFBSs), open chromatin, FANTOM4 enhancers and FANTOM5 enhancers, etc. Genome build 37 coordinates were utilized for all the analyses.

### Asthma phenotyping

Of the total list of available clinical markers, exclusions for markers were made if there was no variation amongst participants. In total, 35 clinical markers were selected to be included in the principal component analysis. If an individual themselves had more than 6 missing values they were excluded from the analysis as well, which resulted in removing 4 individuals leaving a total of 319 to include. 10 of the 35 markers contained missing values following these exclusion processes. For these missing values, the R package mice (v3.16.0) was utilized to impute the missing data. The default settings were utilized, 5 imputations per marker, predictive mean matching (pmm) for continuous traits, logistic regression (logreg) for categorical traits. Distribution plots for these markers prior to and after imputation can be found in Supplemental Figure 1.

### Principal component analysis

Following imputation, the data was normalized using the scale function in R, which performs a z-score transformation for each marker. We then performed principal component analysis (PCA) including 10 principal components (PCs) on our 35 clinical markers that were included. PCA was performed using the robpca() r function via the R package rospca (v1.0.4), which is suited for non-normally distributed data [26, 61]. A scree plot of the eigen values for each of the 10 PCs can be found in Supplemental Figure 7. Outliers for each PC were identified via the R package ewaff (v0.0.2) using the ewaff.handle.outliers function with the following settings: method set to iqr, iqr range set to 3. Outliers were found in PCs 7-10. Principal component 7 contained four outliers, which were removed prior to further analyses (n = 315). Principal components 8-10 each contained several outliers. Due to the nature of the clinical marker data and the way the PCA captured the variance, these PCs largely captured variance from markers recording allergen responses, which resulted in individuals who had strong allergen responses having extreme PC scores. Due to the biological significance of these people, we decided to keep these individuals in the subsequent analyses by dichotomizing the group based on their outlier status (1 = outliers, 0 = not outlier), rather than excluding them. These new dichotomized values for PCs 8, 9, and 10 were then used for the downstream analyses. Frequencies for these PCs can be found in Supplemental Table 11.

### EWAS

Ten epigenome-wide association studies (EWAS) were then performed via the PC scores for each individual for each of these 10 PCs using DNA methylation beta value as the outcome and the PC scores for each individual as the predictor. Due to the related nature of the samples utilized here, each EWAS was conducted via the r package gee (v4.13-25) using a generalized estimating equations (GEE) model that corrected for the correlation structure in families. The following settings were utilized: Gaussian link function, 100 iterations, and the “exchangeable” option to correct for the familial structure. Additionally, each EWAS included age, sex, methylation array row, bisulfite sample plate (dummy-coded), smoking status (codes as 0 = Never smoker, 1 = former smoker, 2 = current smoker), estimated cellular proportions (natural killer cells, monocytes, B cells, CD4+ T cells, CD8+T cells), and lung medication status via questionnaire (inhaled corticosteroids, betamimetica, general lung medication) as covariates. For each EWAS, a standard Bonferroni correction (α = 0.05 / # of CpGs (742,442)) was applied to account for multiple testing to identify significantly associated CpGs.

Enrichment analysis was performed in the EWAS Atlas with the EWAS Toolkit, which compares against previously known associations from previous studies investigating DNA methylation. We also conducted an enrichment analysis via the online tool eFORGE, which is a database of known CpG associations to cell-type specific regulatory elements. This analysis was performed using the “Consolidated Roadmap Epigenomics – Chromatin – All 15-state marks” data set and the standard settings. Next, we cross-analyzed our significantly associated CpGs with significantly associated SNPs (p< 5 × 10^-8^, N= 892 SNPs) identified by Demenais et al in a large meta-analysis [10], utilizing leave-one-out GWAS summary statistics without NTR participants. As has been previously utilized in other studies, a 1 MB range around each CpG was utilized as a cutoff point to identify SNPs within close proximity [62]. Finally, we investigated the CpGs that were found to be within close proximity to a known significantly associated SNP for any previously identified mQTL association by querying the BBMRI mQTL database (based on a previous study by Bonder et. Al.) [27].

Of the 319 individuals included in timepoint 1, 182 had a follow-up blood sample with methylation data available for subsequent analyses at a later time point. For this second timepoint, an additional 10 EWASs were performed using the same PC scores generated from the timepoint 1 clinical marker data to assess longitudinal persistence of the DNA methylation signature associated with these asthma markers. The EWASs were performed with largely the same models as at timepoint 1 samples (stated above) but utilizing covariate data from time point 2. Due to the reduction of individuals and a lack of unique medication covariate combinations, asthma medication status was reduced to a Bayesian variable grouping individuals on the basis of using any one of the three listed medications. This single covariate capturing asthma medication status was then used for these timepoint 2 EWASs. An additional longitudinal comparison was made looking for CpGs in timepoint 1 that were within 100 KB (upstream and downstream) of a CpG identified in timepoint 2. Similar to timepoint 1, following this the timepoint 2 CpGs were grouped and a query of the EWAS Atlas was performed looking for previously identified associations.

## Supporting information

Supplementary Figure 5. Enrichment results from the eFORGE database.

Supplementary Table 4. Independent attachment with complete summary statistics for each of the 10 EWASs.

Supplementary Table 3. Independent attachment with a complete list of significant CpGs for all 10 timepoint 1 EWASs.

## Data Availability Statement

All NTR data can be requested from bona fide researchers (https://ntr-data-request.psy.vu.nl/). Because of the consent given by the study participants, the data cannot be made publicly available. The pipeline for DNA methylation–array analysis developed by the Biobank-based Integrative Omics Study (BIOS) consortium is available at https://molepi.github.io/DNAmArray_workflow/ (https://doi.org/10.5281/zenodo.3355292).

## Acknowledgments

We would like to acknowledge all the participants in the Netherlands Twin Register for their willingness to participate in this study. We would also like to acknowledge the BIOS Consortium. A list of the contributors to this consortium and other additional details can be found at https://www.bbmri.nl/bios-consortium.

## Supplementary Tables and Figures

**Supplementary Table 1.** Table showing PC eigen values and % of total variance captured.

| PC | Eigen Value | % Variance | Cumulative % |
| --- | --- | --- | --- |
| PC1 | 4.4832 | 35.27% | 35.27% |
| PC2 | 2.4844 | 19.54% | 54.81% |
| PC3 | 1.5834 | 12.46% | 67.26% |
| PC4 | 1.2396 | 9.75% | 77.01% |
| PC5 | 1.0671 | 8.39% | 85.41% |
| PC6 | 0.7696 | 6.05% | 91.46% |
| PC7 | 0.5435 | 4.28% | 95.74% |
| PC8 | 0.2794 | 2.20% | 97.94% |
| PC9 | 0.1555 | 1.22% | 99.16% |
| PC10 | 0.1071 | 0.84% | 100.00% |
| Total | 12.7125 |  |  |

**Supplementary Table 2.** EWAS results for time point 1 with inflation values.

| PC | Inflation |
| --- | --- |
| PC1 | 0.999737038 |
| PC2 | 0.894699702 |
| PC3 | 0.861547269 |
| PC4 | 1.034053431 |
| PC5 | 1.021476296 |
| PC6 | 0.749147555 |
| PC7 | 0.805103148 |
| PC8 | 1.003658406 |
| PC9 | 0.843756081 |
| PC10 | 0.848921965 |

**Supplementary Table 3.** Independent attachment with a complete list of significant CpGs for all 10 timepoint 1 EWASs.

**Supplementary Table 4.** Independent attachment with complete summary statistics for each of the 10 EWASs.

**Supplementary Table 5.**
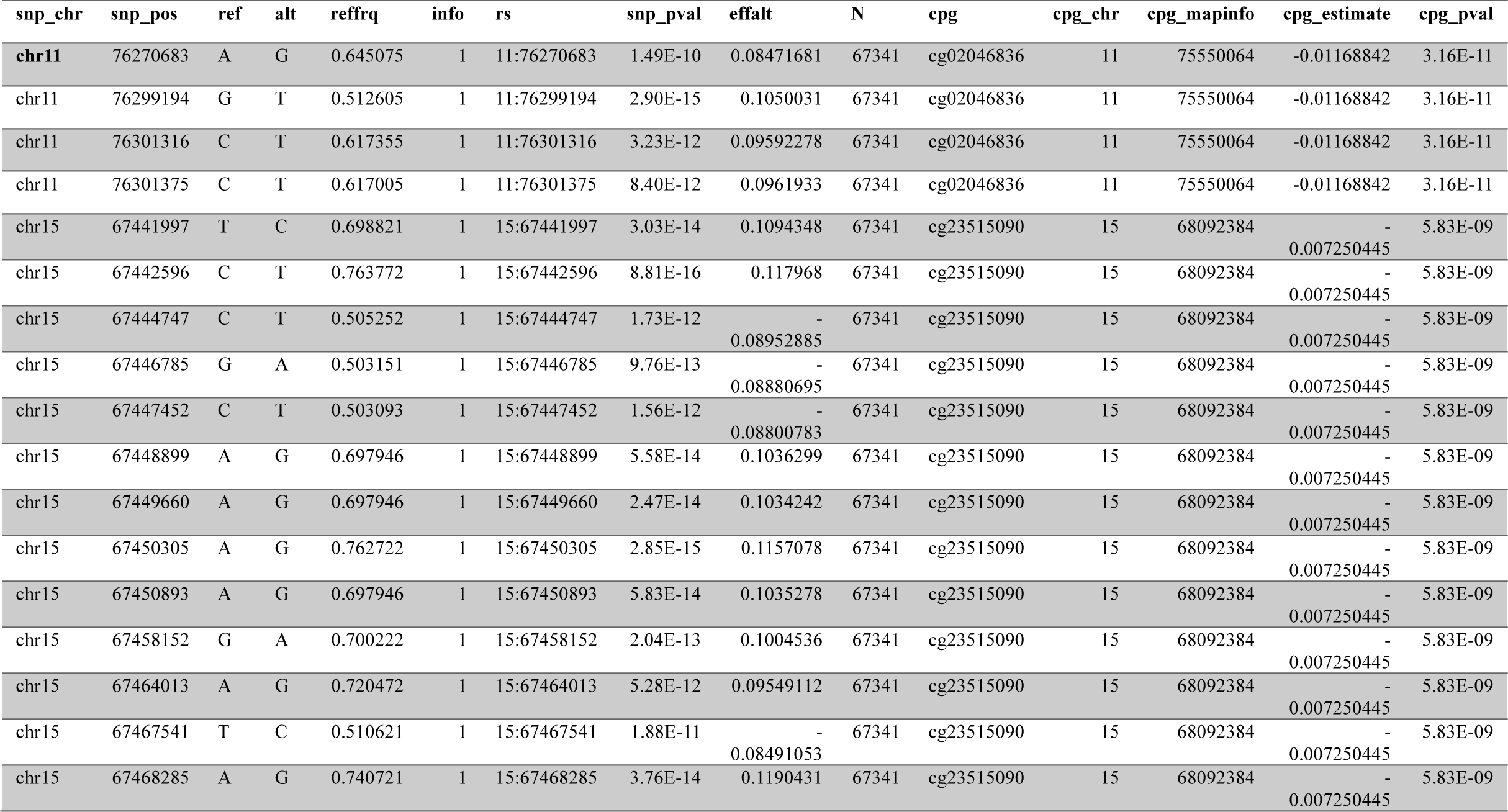
Summary table of the GWAS SNPs identified by Demenais et. al. to reside within 1 MB of a CpG identified at timepoint 1.

**Supplementary Table 6.** Independent attachment with a complete list of significant CpGs for all 10 timepoint 2 EWASs.

**Supplementary Table 7.** EWAS results for time point 2 with inflation values.

| PC | Inflation |
| --- | --- |
| PC1 | 1.027943459 |
| PC2 | 0.979550307 |
| PC3 | 0.999929682 |
| PC4 | 1.000268796 |
| PC5 | 0.775029681 |
| PC6 | 0.949333881 |
| PC7 | 0.952948417 |
| PC8 | 1.004404304 |
| PC9 | 0.999975322 |
| PC10 | 1.02109527 |

**Supplementary Table 8.** The CpGs from timepoint 1 that reside within 100 kb of a CpG identified at timepoint 2.

| Genome_Build | TP1_ilmnID | TP1_CHR | TP1_MAPINFO | TP1_estimate | TP1_pval | TP1_PC | TP2_ilmnID | TP2_CHR | TP2_MAPINFO | TP2_estimate | TP2_pval | TP2_PC |
| --- | --- | --- | --- | --- | --- | --- | --- | --- | --- | --- | --- | --- |
| 37 | cg01043901 | 13 | 43061678 | -0.0061855 | 1.23E-08 | PC7 | cg04217496 | 13 | 43119835 | 0.003371794 | 4.51E-08 | PC7 |
| 37 | cg04983687 | 16 | 88558223 | -0.0269112 | 1.64E-12 | PC7 | cg09247061 | 16 | 88571844 | 0.005722604 | 2.14E-08 | PC2 |
| 37 | cg08940169 | 16 | 88540241 | -0.0130795 | 2.54E-09 | PC7 | cg09247061 | 16 | 88571844 | 0.005722604 | 2.14E-08 | PC2 |
| 37 | cg12077754 | 2 | 75089669 | -0.0128193 | 2.36E-08 | PC7 | cg19359099 | 2 | 75060770 | -0.000276636 | 1.97E-09 | PC3 |

**Supplementary Table 9.** Questionnaire responses for two surveys asking about asthma status prior to study enrollment for the initial 425 individuals.

|  | Survey 1 | Survey 2 |
| --- | --- | --- |
| **No** | 207 (65.1%) | 219 (61.7%) |
| **Yes** | 111 (34.9%) | 136 (38.3%) |
| **Total** | 318 | 355 |
| **Response Rate (%)** | 74.8% | 83.5% |

**Supplementary Table 10.**
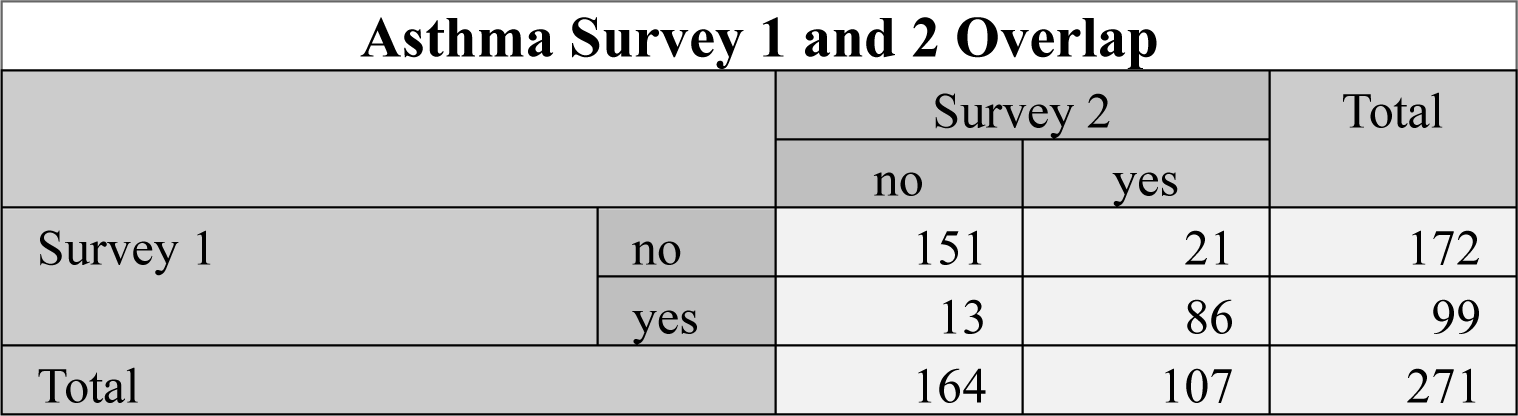
Comparison of survey responses from individuals who responded to both surveys.

| Asthma Survey 1 and 2 Overlap |  |  |  |  |
| --- | --- | --- | --- | --- |
|  |  | Survey 2 |  | Total |
|  |  | no | yes |  |
| Survey 1 | no | 151 | 21 | 172 |
|  | yes | 13 | 86 | 99 |
| Total |  | 164 | 107 | 271 |

**Supplementary Table 11.** Frequencies of outliers for principal components 8, 9, and 10. Outlier status was determined using thresholds of Q1 – (3 x IQR) and Q3 + (3 x IQR).

| PC | Non-outliers | Outliers |
| --- | --- | --- |
| **PC8** | 265 | 54 |
| **PC9** | 266 | 53 |
| **PC10** | 272 | 47 |

**Supplementary Figure 1.**
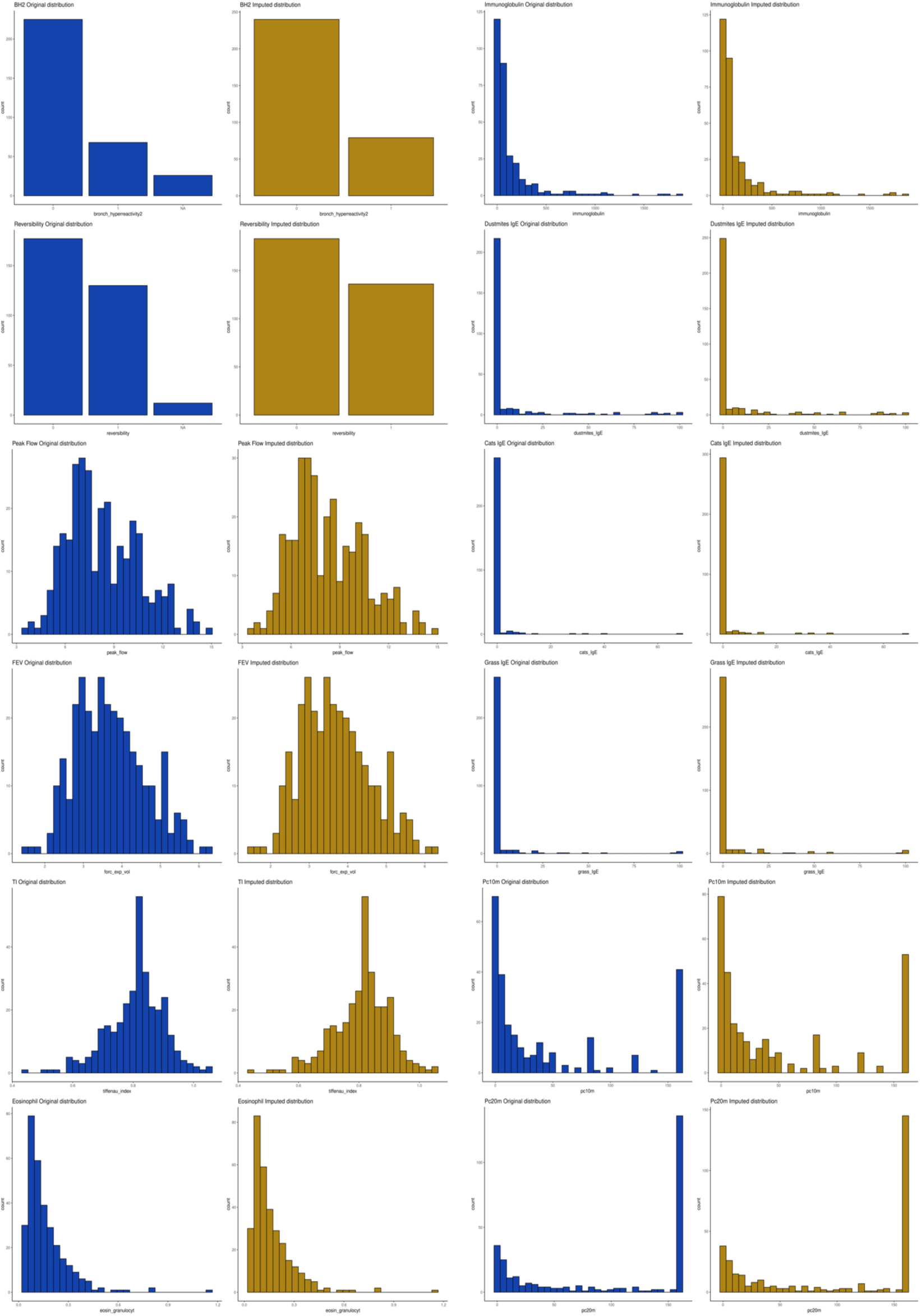
Distribution plots of the imputed clinical markers of asthma prior to (blue) and after imputation (yellow).

**Supplementary Figure 2.**
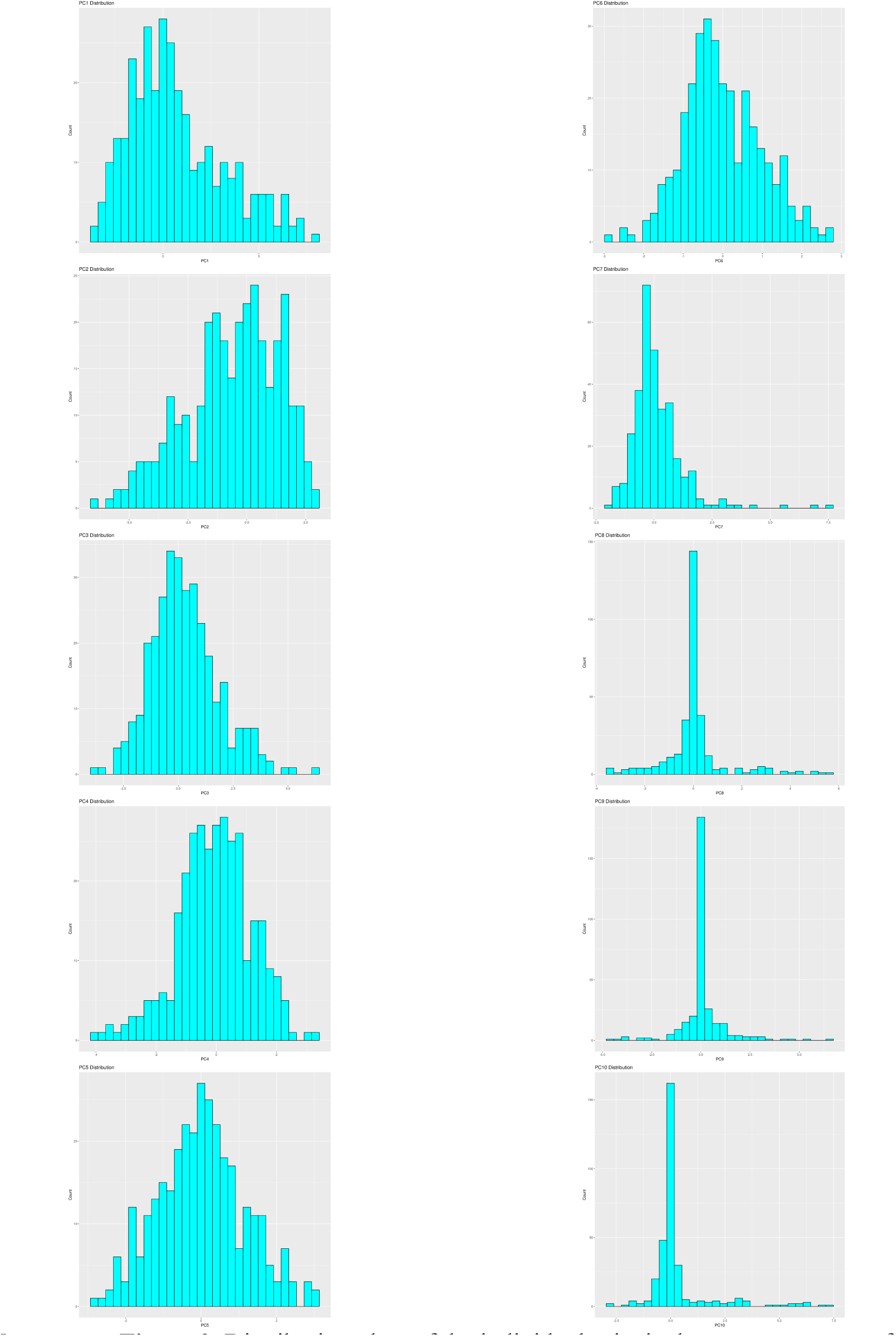
Distribution plots of the individual principal component scores for each PC.

**Supplementary Figure 3.**
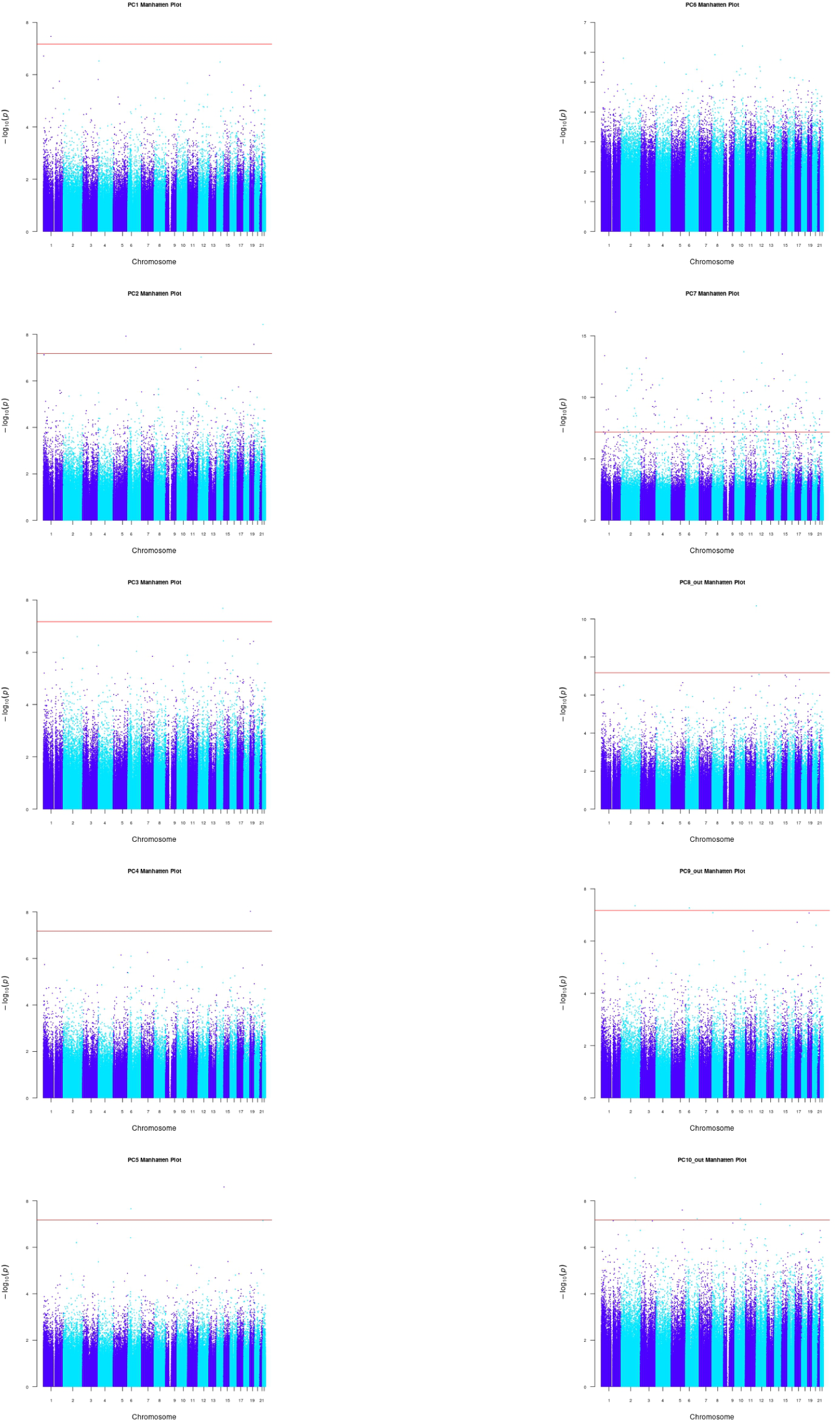
EWAS results calculated the 10 different sets of PC scores from the first timepoint. The red line indicates the significance threshold set via a Bonferroni correction.

**Supplementary Figure 4.**
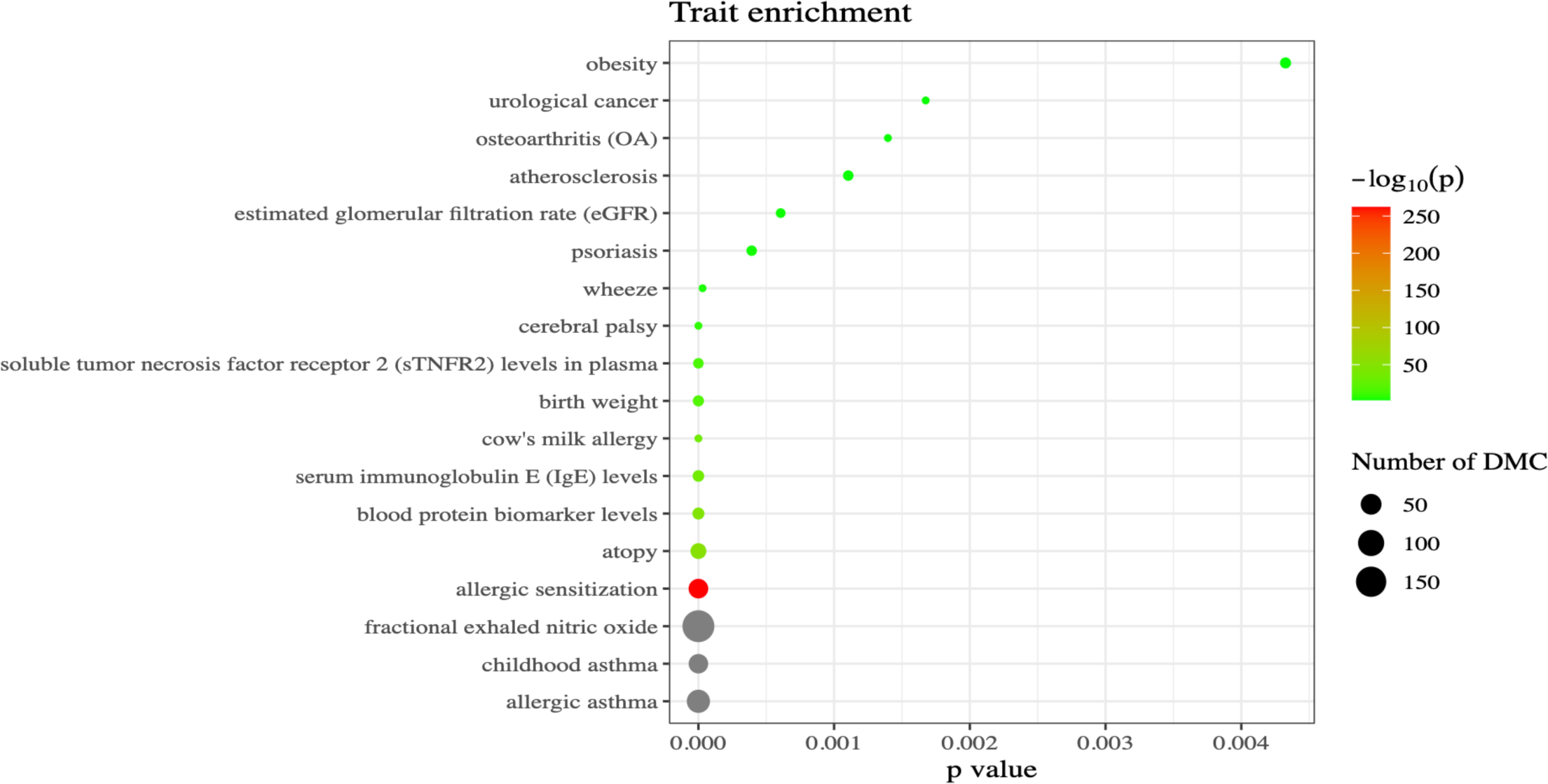
EWAS atlas enrichment analysis of the 204 CpGs identified at TP1 from PC7. The EWAS Atlas job can be found via this job ID: 2a37ab12d2785eaa0d564e062cff5f96.

**Supplementary Figure 5.**
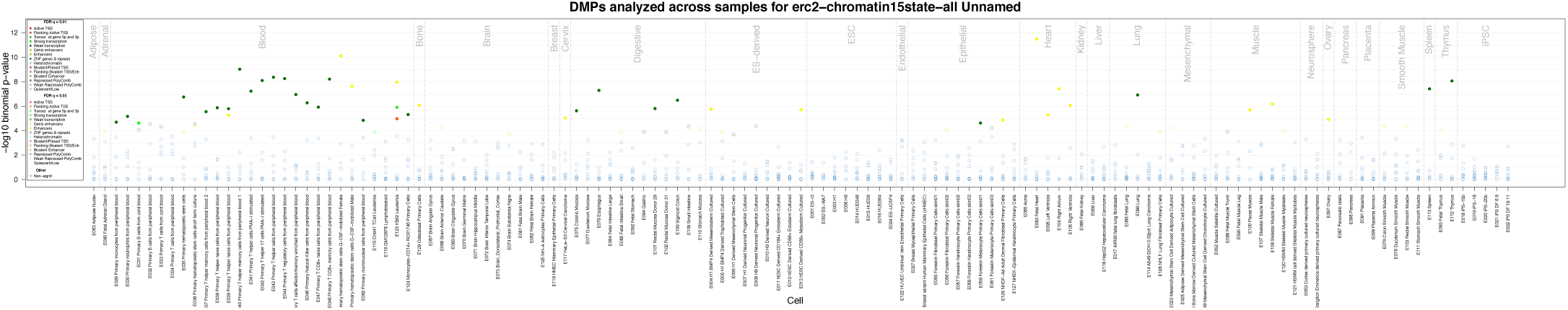
Enrichment results from the eFORGE database (separate PDF included for better visualization).

**Supplementary Figure 6.**
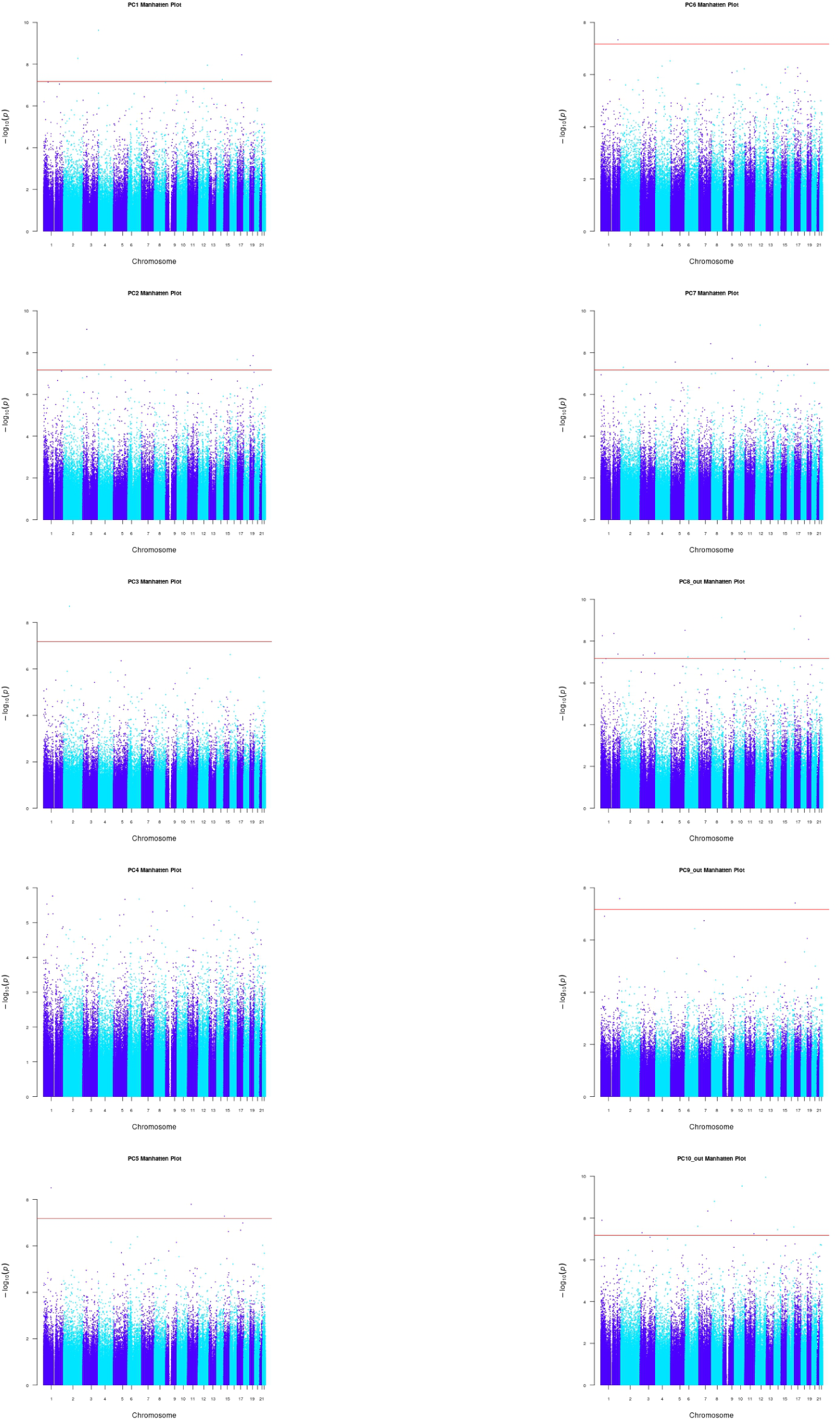
EWAS results calculated the 10 different sets of PC scores from the second timepoint. The red line indicates the significance threshold set via a Bonferroni correction.

**Supplementary Figure 7.**
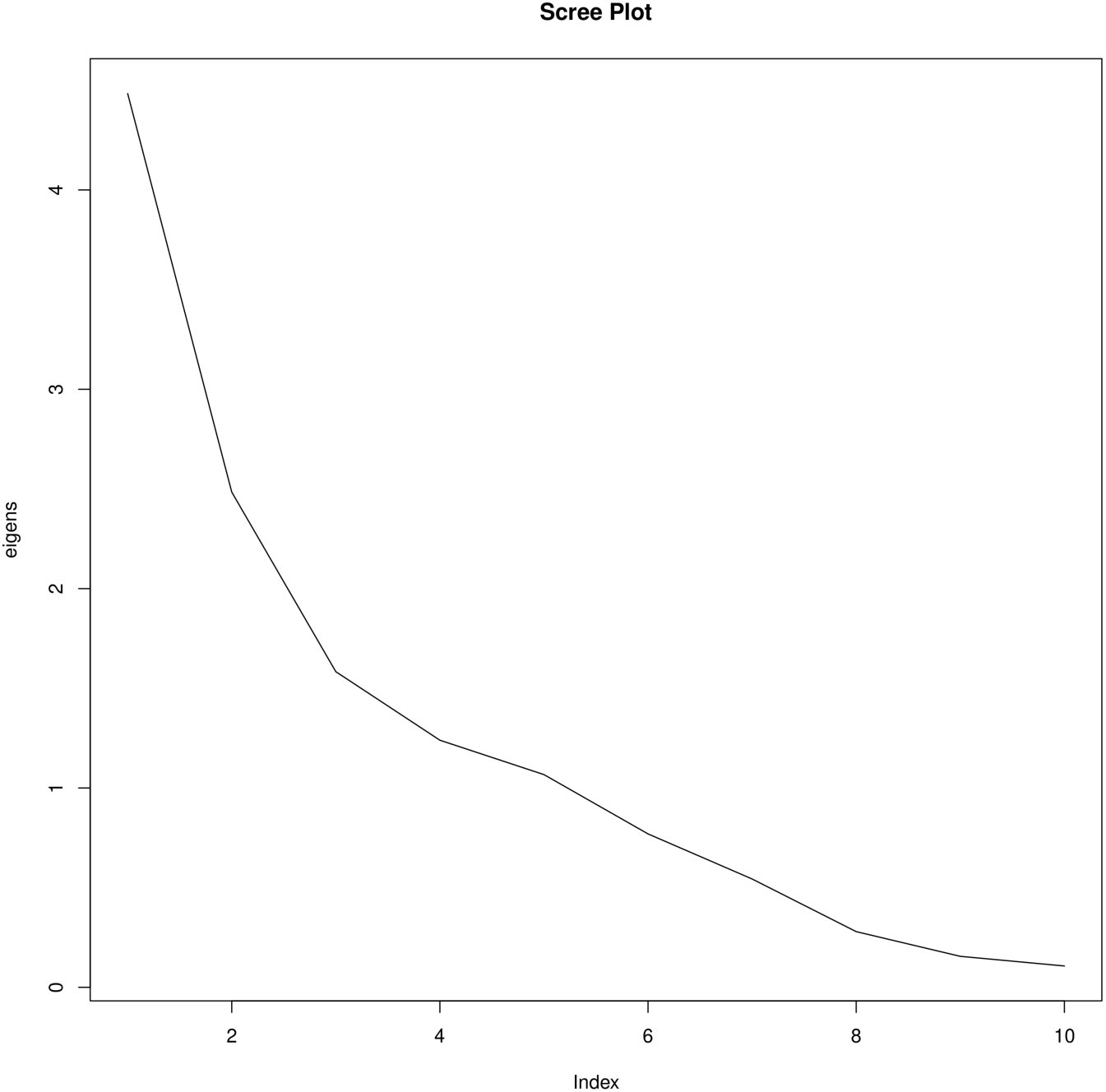
Scree plot showing the raw eigen values for each of the 10 principal components included.

