## Supplementary figures and images for "Epigenetic Signatures of Asthma: A Comprehensive Study of DNA Methylation and Clinical Markers"

### Supplementary Figure 5. Enrichment results from the eFORGE database.

#### DMPs analyzed across samples for erc2−chromatin15state−all Unnamed

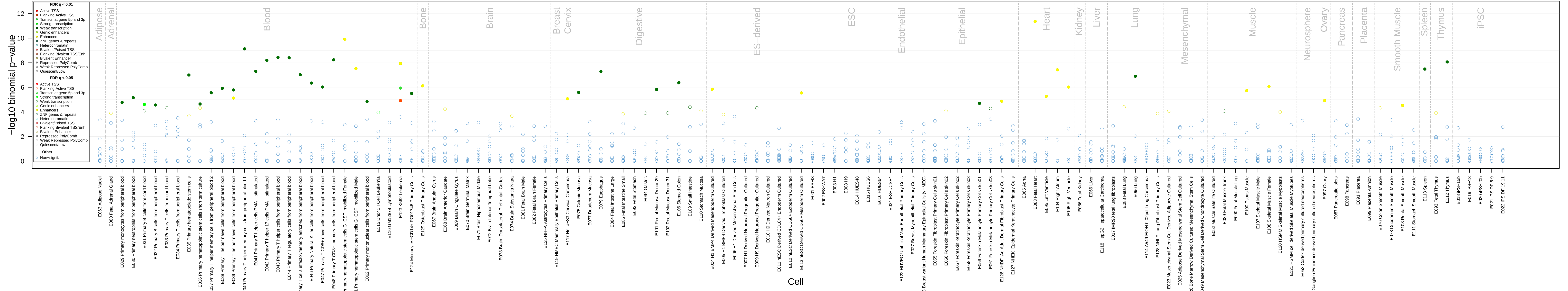
